# Post-stroke reorganization of transient brain activity characterizes deficits and recovery of cognitive functions

**DOI:** 10.1101/2021.03.16.21253745

**Authors:** Elvira Pirondini, Nawal Kinany, Cécile Le Sueur, Joseph C. Griffis, Gordon L. Shulman, Maurizio Corbetta, Dimitri Van De Ville

## Abstract

Functional magnetic resonance imaging (fMRI) has been widely employed to study stroke pathophysiology. In particular, analyses of fMRI signals at rest were directed at quantifying the impact of stroke on spatial features of brain networks. However, brain networks have intrinsic time features that were, so far, disregarded in these analyses. In consequence, standard fMRI analysis failed to capture temporal imbalance resulting from stroke lesions, hence restricting their ability to reveal the interdependent pathological changes in structural and temporal network features following stroke. Here, we longitudinally analyzed hemodynamic-informed transient activity in a large cohort of stroke patients (n = 103) to assess spatial and temporal changes of brain networks after stroke. While large-scale spatial patterns of these networks were preserved after stroke, their durations were altered, with stroke subjects exhibiting a varied pattern of longer and shorter network activations compared to healthy individuals. These temporal alterations were associated with white matter damage and were behavior-specific. Specifically, restoration of healthy brain dynamics paralleled recovery of cognitive functions, but was not significantly correlated to motor recovery. These findings underscore the critical importance of network temporal properties in dissecting the pathophysiology of brain changes after stroke, thus shedding new light on the clinical potential of time-resolved methods for fMRI analysis.

**Significance Statement:** Understanding the pathophysiology of a disorder is pivotal to design effective treatment. In this regard, recent advances in stroke research settled a new clinical concept: *connectional diaschisis*, which suggested that post-stroke impairments arise from both focal structural changes (tied to the injury) and widespread alterations in functional connectivity. fMRI time-resolved methods consider structural and temporal properties of brain networks as interdependent features. They are, thus, better suited to capture the intertwine between structural and functional changes. Here we leveraged a dynamic functional connectivity framework based on the clustering of hemodynamic-informed transients in a large and heterogeneous stroke population assessed longitudinally. We showed that lesions led to an unbalance in the brain dynamics that was associated with white matter fibers disruption and was restored as deficits recovered. Our work showed the potential of a time-resolved method to reveal clinically relevant dynamics of large-scale brain networks.

## Introduction

The mammalian brain, even in the absence of explicit task, operates through the continuous integration and segregation of signals from different brain areas. Since the landmark work from Fox and colleagues in 2005 (1), functional magnetic resonance imaging (fMRI) performed at rest has become one of the most prominent methods to investigate intrinsic brain activity and its relationship with behavior or psychopathology (2–4). These task-free resting-state paradigms could potentially be advantageous to measure pathological brain changes, as they can be readily deployed, even with patients unable to match control performance due to motor and cognitive impairment (5–7)3/16/21 3:10:00 PM.

Analysis of resting-state fMRI (rs-fMRI) has so far mostly relied on measuring inter-regional (or voxel- or vertex-level) connectivity by means of Pearson correlation between time-series from a set of predefined regions of interest (a.k.a. *static* functional connectivity). In these studies, information exchange between neuronal populations of different regions is assumed to engender stronger statistical dependency stationary over time. Nevertheless, the human brain is a dynamic system that fluctuates at the time scale of seconds (2). Therefore, *static* connectivity approaches, despite their methodological simplicity and ease, may miss features reflecting the inherent dynamic nature of the brain. In the last decade, several time-resolved approaches have thus been proposed to investigate the so-called *dynamic* functional connectivity (dFC) ((8) for a review). They have been demonstrated to provide benefits over *static* methods, notably to study cognition and psychiatric disorders (4, 9–11). Besides, deeming non-stationarity enables a more accurate description of the modular interactions occurring between brain functional networks and their anatomical substrate (12).

Despite this potential, only a handful of studies have employed time-tailored methods to explore the neural correlates of stroke (13–17), even though it is one of the major neurological disorders in Western societies and a leading cause of long-term disabilities. Ranging from motor to cognitive deficits, these disabilities arise from both focal structural changes (tied to the injury) and widespread functional alterations in inter-regional connectivity (18, 19), as theorized under the concept of *connectional diaschisis* (20). Structural and functional abnormalities combine in an interdependent manner to generate both deficits and recovery processes. Considering the complexity of these interactions, time-resolved FC approaches, which spatial *and* temporal properties of brain networks, could help unravel the intertwine between structural disruptions and lesion-induced dynamic changes in large-scale functional networks. Coupled to behavioral and clinical assessments, these methods could further elucidate the nature of pathological changes occurring after stroke, possibly supporting our understanding of recovery processes (21, 22).

Here, we leveraged a recent dynamic FC framework, i.e., the innovation-driven coactivation patterns (iCAP) framework (1, 2), to investigate whether spatial and temporal properties of large-scale brain networks following stroke correlate with anatomical damage and behavioral recovery. We applied the iCAP framework to an extensive dataset (n = 103) of stroke patients scanned at different points in time after-lesion (i.e., 1-2 weeks, 3 months and 1 year) and with different neurological syndromes (i.e., ranging from motor to cognitive deficits). While the spatial patterns of the obtained large-scale functional brain networks were preserved after stroke, lesions disrupted their temporal durations, emphasizing the prospects of exploiting time-resolved fMRI methods. Importantly, these altered durations were *i)* proportional to the percentage of disrupted white matter fibers; *ii)* specific to the neuropsychological deficit; iii) correlated with functional improvements; and *iv)* restored over time proportionally to the recovery of deficits.

This is the first time that a time-resolved approach is deployed in a large cohort of stroke patients, which were evaluated longitudinally with a comprehensive set of multi-domain clinical assessments. Using the iCAP framework, we revealed important aspects of post-lesional reorganization of functional dynamics, in relation to anatomy and to behavioral changes. We posit that understanding the nature of this relationship is pivotal to grasp the multifaceted reorganization mechanisms that occur following stroke, and that are directly involved in recovery (19, 21, 27, 28).

## Results

### Lesion topography and behavioral profile of patients

We studied a large population of stroke patients and age-matched healthy subjects (i.e., in total 267 acquisitions, **Table S1**). Briefly, first-time stroke patients with clinical evidence of impairment were recorded longitudinally: 1-2 weeks (n = 103), 3 months (n = 72), and 12 months (n = 54) after the lesion. The healthy control group (n = 19) also underwent two imaging sessions at a distance of 3 months. Lesion volume varied greatly over patients (ranging from 0.1 cm^3^ to 277.02 cm^3^, mean ± SD volume: 33.66 cm^3^ ± 49.51 cm^3^), with the highest overlap found in white matter and subcortical regions (**Figure S1a**). Specifically, the most affected tracts were the corticospinal tract, the fronto- and parieto-pontine tract, the extreme Capsule (EMC), and the inferior fronto-occipital fasciculus (IFOF) (**Figure S1b-c**). Prior to each fMRI scan, patients were tested with an exhaustive neurobehavioral battery. We applied principal component analysis (PCA) on these behavioral data to isolate clusters of deficits for each domain separately (e.g., motor, language, attention, spatial memory, and verbal memory) (29). We found factors analogous to our previous works (see **SI** (29, 30)) and also similar recovery effects (30, 31). Indeed, patients with severe acute deficits (i.e., > 2 SD from healthy subjects at 1-2 weeks post-lesion) recovered mostly within the first three months post-lesion (**Figure S1d**). Instead, patients with less severe acute deficits (i.e., < 2 SD from healthy subjects) had stable performances over sessions comparable to those of the healthy subjects.

### Whole brain dynamics is preserved in patients

We applied Total Activation on the denoised BOLD time courses to retrieve robust transient activity (i.e., frames with highly changing activity, the so-called significant innovation frames), individually for the 267 acquisitions considered (**Figure S2**). Patients and healthy subjects had a comparable number of transients (mean ± SD - healthy controls: 43.8 ± 12.0; patients 1-2 weeks: 45.8 ± 22.4; 3 months: 43.4 ± 19.5; 1 year: 43.7 ± 20.4, expressed in percentage of the total number of frames for each subject - **Figure 1a**), highlighting that the whole brain overall amount of dynamic fluctuations was preserved after stroke. Yet, dynamics of specific brain regions could still be affected by the lesion. For this reason, we then extracted spatial patterns of large-scale networks and we computed dynamic features for each network individually.

**Figure 1.**
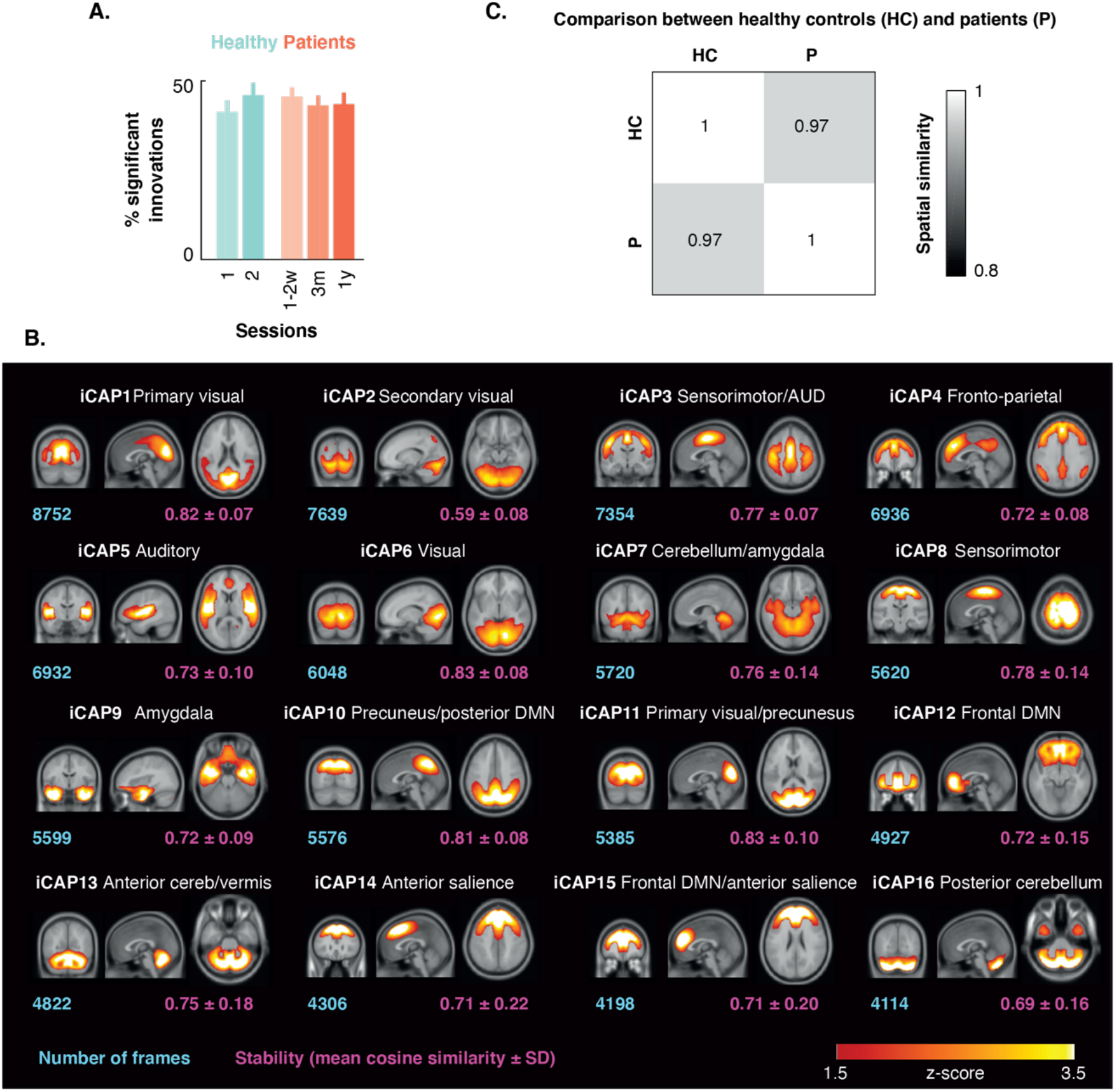
iCAP networks and spatial features. **A**. Number of transients (or significant innovation frames) including both positive and negative frames expressed in percentage of the total number of frames for the two sessions of the healthy control subjects (in cyan) and for the three sessions of the stroke patients (in orange). **B**. Spatial patterns (displayed in Montreal Neurological Institute coordinates) for the 16 innovation-driven coactivation patterns (iCAPs) retrieved from all subjects, including both sessions of the healthy control subjects and the three sessions of the stroke patients. Blue values denote the total number of significant innovation frames of each cluster (i.e., iCAP) and purple values indicate the stability of each cluster calculated as the mean cosine similarity from the centroid over subjects (mean ± SD over subjects). AUD, auditory; DMN, default mode network; cereb, cerebellum. **C**. Cosine similarity between iCAP spatial patterns for healthy controls (HC) and patients (P) averaged over the 16 iCAPs.

### Resting-state activity can be decomposed into spatial maps corresponding to known functional networks

Transients of both healthy subjects and patients at different time points were fed to a k-means clustering to obtain stable innovation-driven co-activation patterns (iCAPs), which could be spatially and temporally overlapping. The number of clusters was determined using evidence accumulation and, as a result, 16 iCAPs were extracted (**Figure S3a**). The iCAPs corresponded to well-known resting-state networks, in line with our previous results in different datasets (**Figure 1b** and **Table S2**) (23–25). Specifically, they included sensory-related networks such as primary and secondary visual areas (iCAP 6), auditory and language network (iCAP 5), with high activations in Heschl gyrus and rolandic operculum, and sensorimotor network (iCAPs 3 and 8), which included pre-, post-, and para-central areas, supplementary motor area, and middle cingulum (only for iCAP 3). Two iCAPs (1 and 11) represented the visuospatial/ventral attention network and comprised the primary (iCAP 1) and secondary (iCAP 11) visual areas along with the precuneus. There was an additional task-related iCAP, the fronto-parietal network (iCAP 4), with high activations in the frontal and parietal lobes together with anterior and middle cingulate cortex (ACC and MCC). As previously reported (23–25), the Default Mode Network (DMN) was decomposed into a frontal part (iCAP 12) including frontal lobe and ACC, and a posterior part (iCAP 10) comprising the cuneus, the precuneus, and the superior and inferior parietal lobes. Four iCAPs included regions of the cerebellum (iCAPs 2, 7, 13, and 16). Yet, iCAPs 13 and 16 were specific to the anterior (iCAP 13) and posterior cerebellum (iCAP 16); whereas iCAP 2 included as well secondary visual areas and iCAP 7 hippocampal areas. The remaining iCAPs comprised regions of the salience network, with iCAP 15 encompassing middle and superior frontal lobe and ACC, and iCAP 9 related to the inferior temporal lobe, the hippocampus and the amygdala.

### iCAP spatial maps are similar between patients and controls

Prior to evaluating differences between patients and healthy controls, we first assessed between-session spatial replicability of the iCAPs in healthy subjects. The high similarity of the spatial patterns (mean cosine similarity over iCAPs ± SD: 0.95 ± 0.02 **Figure S4a**) emphasized the ability of the iCAP framework to extract reliable and stable networks. Interestingly, spatial maps were also very similar between patients and healthy controls (mean cosine similarity over iCAPs ± SD at 2 weeks: 0.97 ± 0.03; 3 months: 0.97 ± 0.02; 1 year: 0.97 ± 0.01 - **Figure 1c** and **Figure S4b**), as previously reported using static functional connectivity (27). Finally, spatial maps were also highly comparable between left and right damage patients (**Figure S4c**).

### Patients exhibited a varied pattern of longer and shorter resting-state network activations

The stability of the spatial maps over sessions (**Figure S4a-c**) and groups (**Figure 1c**) highlighted the robustness of the brain’s functional architecture, as described using iCAPs. We capitalized on this observation to investigate the temporal properties associated with these large-scale brain networks. To this end, we used spatiotemporal transient-informed regression to derive iCAP time courses at the subject-level and to compute the average duration of each iCAP. Similarly to our assessment of spatial replicability, we evaluated between-session temporal replicability in the healthy subjects. Given the stability of average iCAP durations in the healthy control group (p > 0.31 - Bonferroni-Holm FWE corrected; see **Figure S4d**), we then probed whether this temporal measure could be leveraged as a discriminative feature to distinguish stroke patients and healthy subjects. Considering that patients present with a combination of deficits in different domains, which may reflect dysfunction across multiple brain networks (7), we opted for a multivariate approach, which considers a combination of multiple features instead of individual networks (32). Specifically, after selecting the six most stable iCAPs over folds of patients and healthy subjects using a two-sample Mann-Whitney test, we performed a Linear Discriminant Analysis (LDA) (33) that allowed to discriminate between the two groups with a significant accuracy (0.66 ± 0.04 - p < 0.02), as illustrated by a diagonal confusion matrix (**Figure 2a**), and sensitivity-specificity, as emphasized by the area-under the curve (AUC: 0.63 – null distribution threshold: 0.60). LDA weights were highly stable over cross-validation folds (mean absolute z-score ± SD over iCAPs: 15.49 ± 6.46) and captured specific regions whose temporal profiles were affected in stroke patients. In particular, patients showed a longer duration in pre-central areas, anterior and medium cingulum, and superior temporal lobe. Instead, the occipital lobe, inferior temporal lobe and the postero-lateral cerebellum had a reduced duration (**Figure 2b-c**). A similar varied pattern of longer and shorter network activations was observed when comparing individual iCAP durations between healthy controls and stroke patients at different point in time after lesion (**Figure S4e**).

**Figure 2.**
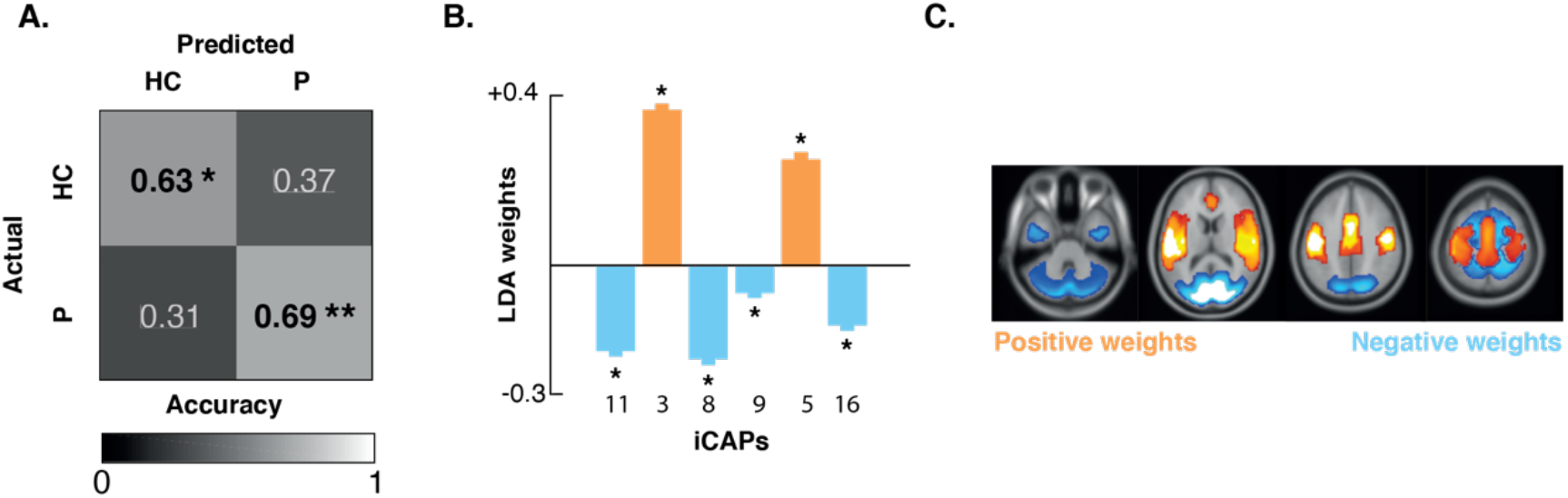
Temporal properties: comparison between healthy controls and patients. **A**. Confusion matrix for patients and healthy controls for the Linear Discriminant Analysis classifier built using the iCAP durations of the 6 most stable iCAPs. * p < 0.05; ** p < 0.01 (non-parametric permutation testing). **B**. LDA weights for the 6 most stable iCAPs (mean ± SD over folds). * indicate significant values - i.e., weights with mean/SD over cross-validation folds > 3.2. **C**. Linear combination of the 6 most stable iCAP spatial patterns weighted by the LDA weights displayed in Montreal Neurological Institute coordinates. Blue represents locations with shorter duration for stroke patients; whereas orange represents locations with longer duration.

### Changes in duration are explained by disconnections within white-matter tracts

In order to better understand the interplay between functional dynamics and anatomy, we deployed a partial least square correlation (PLSC) analysis between iCAP average durations and tract integrity (**Figure 3a**). We found a single significant component (p-value = 0.003; ρ = 0.35), only for patients at 1-2 weeks post-lesion. The longer duration of the iCAPs (**Figure 3b**) was explained by the loss of white matter fibers, in particular of *i)* the projection pathways, which connect cortical areas with subcortical nuclei and brainstem; *ii)* a few association pathways which connect disparate cortical areas including the cingulum, the EMC, the IFOF, and the uncinate fasciculus; *iii)* the anterior and middle commissure; and *iv)* the fiber bundles inside the brainstem (i.e., the medial lemniscus and the spinothalamic tract). Interestingly, the fiber bundles in the cerebellar pathways, and specifically the inferior cerebellar peduncle had opposite saliences highlighting a different behavior for the cerebellar structure, characterized by a lower average duration as compared to healthy controls.

**Figure 3.**
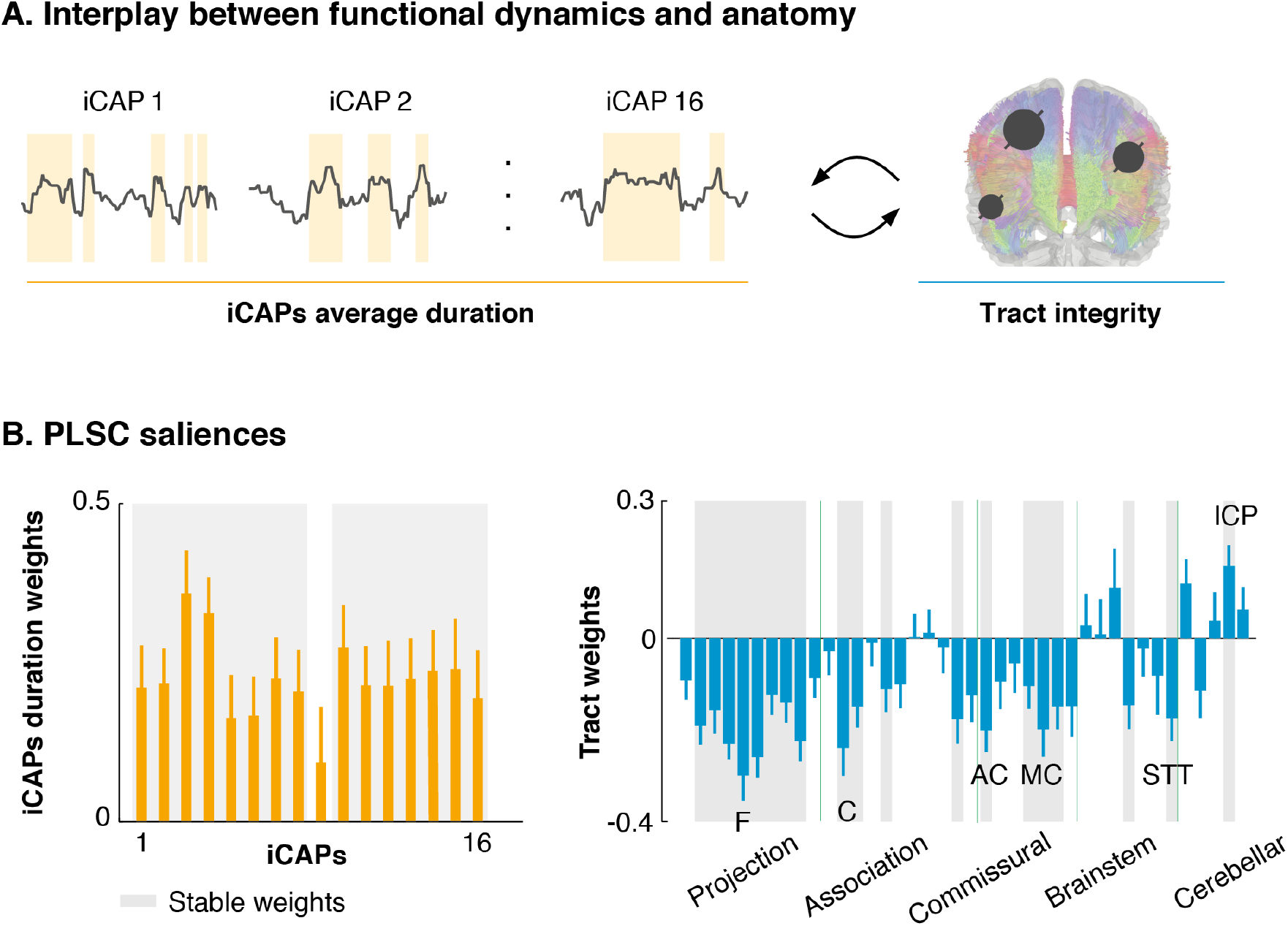
Interplay between functional dynamics and tract integrity. **A**. We deployed a partial least square correlation (PLSC) analysis between iCAP average durations and tract integrity for patients at 2 weeks post-lesion (27, 37). **B**. Left panel: iCAP durations saliences for the significant PLS component (p = 0.003) (mean ± SD over bootstrap repetitions); Right panel: tract integrity saliences of the significant PLS component (mean ± SD over bootstrap repetitions). Tracts were grouped in projection pathways, association pathways, commissural pathways, brainstem pathways, and cerebellar pathways following (42). Grey shadows indicate stable salience, i.e., saliences lower/higher than the lower/upper bound of 95% confidence interval of bootstrapping distributions (bootstrapping procedure with 500 random samples with replacement). F: fornix; C: cingulum; AC: anterior commissure; MC: medial commissure; STT: spinothalamic tract; ICP: inferior cerebellar peduncle.

### Changes in duration are associated with behavioral deficits and recovery

We then assessed whether changes in iCAP durations correlated with clinical impairments and recovery, as estimated using the behavioral measures. We applied a second PLSC analysis, this time between the temporal measures and the behavioral data of the three time points, separately for each domain. Functional network durations showed significant correlations with recovery from attention, language, and spatial memory deficits (attention: p-value = 0.03, ρ = 0.23; language: p-value = 0.003, ρ = 0.18; spatial memory: p-value = 0.007, ρ = 0.18), but not from motor or verbal memory impairments (p-value > 0.12). This observation parallels previous findings regarding the relationship between brain modularity and behavior, in which a reduction in modularity was observed in the sub-acute stage, followed by a partial recovery, limited to attention, language and spatial memory (31). To further explore the clinical relevance of our results, we first investigated the saliences related to the behavioral data. We observed that they were only significant for patients with severe acute deficits, which were the ones showing the largest recovery over time (**Figure S1d**). We then investigated the saliences pertaining to iCAP durations and highlighted that temporal patterns were behavior-specific (mean correlation ± SD over behaviors: 0.22 ± 0.08, **Figure 4a**). Indeed, attention domain had saliences in the fronto-parietal network, the amygdala, the precuneus and the visual cortex. Spatial memory symptoms, instead, were mostly correlating with fronto-parietal and sensorimotor networks, the posterior part of the cerebellum, the amygdala, and the visual cortex. Finally, language-related deficits were specifically tied to the language network and primary and secondary visual areas. In order to probe the temporal evolution of these brain functional components, we then projected the individual subject’s iCAP durations onto the significant multivariate brain saliences for healthy controls and for patients with severe acute deficits (**Figure 4b**). Over time, the brain scores of stroke patients approached those of healthy controls, suggesting that post-stroke network dynamics restored proportionally to behavioral improvement.

**Figure 4.**
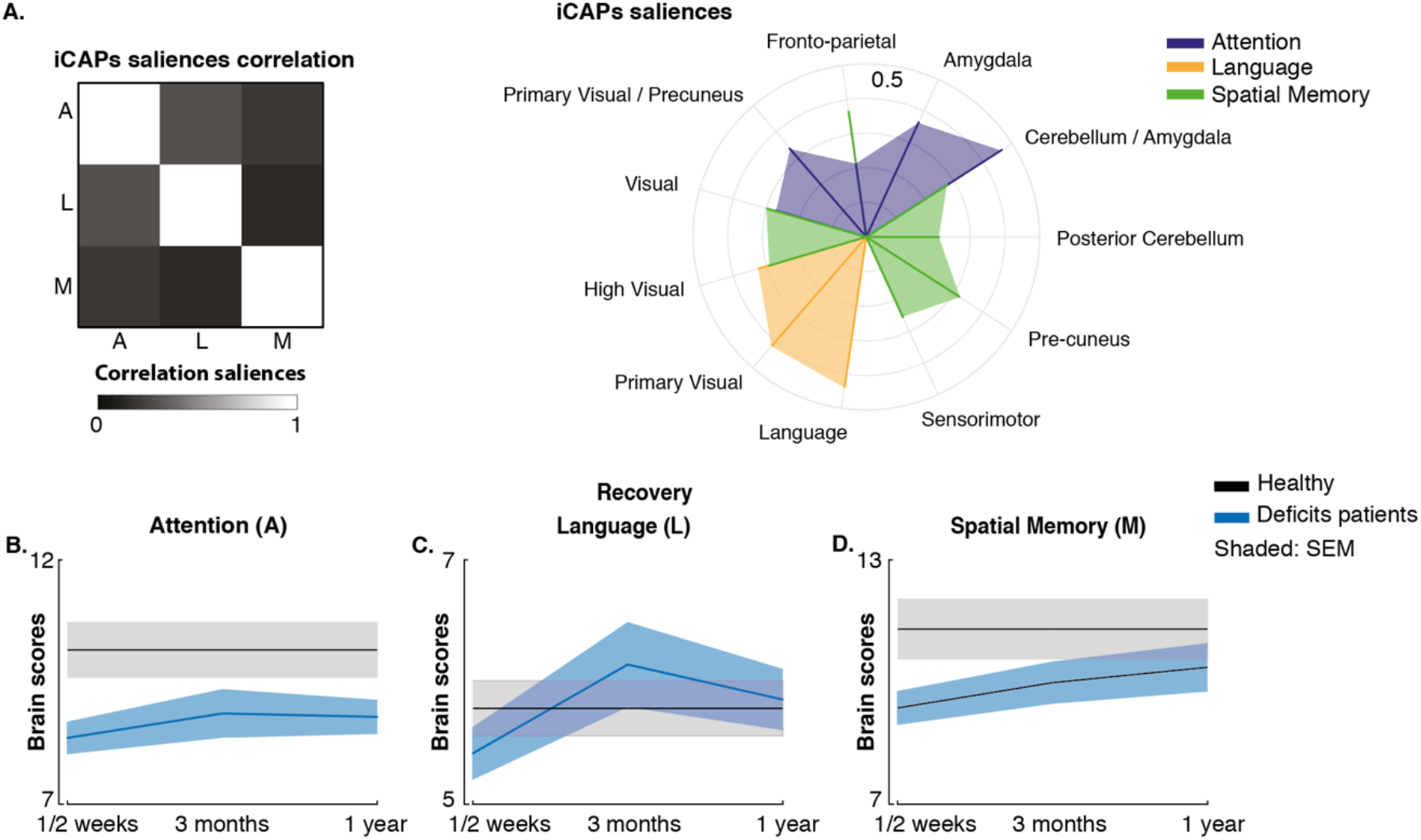
Interplay between functional dynamics and behavioral deficits. We deployed a partial least square correlation (PLSC) analysis between iCAP average durations and behavioral scores and recovery for all patients and all sessions. **A**. Left panel: correlation matrix between iCAP durations saliences between the three behavior domains that had one significant PLS component (i.e., attention, language, and spatial memory). Right panel: iCAP durations saliences for attention (blue), language (orange), and spatial memory (green). **B**. Brain scores obtained projecting the individual subject’s iCAP durations onto the significant (i.e., saliences lower/higher than the lower/upper bound of 95% confidence interval of bootstrapping distributions (bootstrapping procedure with 500 random samples with replacement)) multivariate brain saliences (i.e., **Lx=XV**, see **Partial Least Square section** in **SI**) for healthy controls (black) and for patients with severe acute deficits (blue) for the three behavior domains and the three time points (i.e., 1-2 weeks, 3 months, and 1-year post-lesion) separately. Shadows represent SEM.

## Discussion

In recent years, *dynamic* functional connectivity methods demonstrated to provide additional insights into the rich spatiotemporal orchestration of spontaneous fluctuations as compared to *static* approaches (2, 8–11), in particular for psychiatric conditions (4). However, the transition towards dFC in the context of stroke has so far remained limited (13–17). While existing studies provided compelling evidence in favor of a dynamic reconfiguration of brain networks following stroke, they relied on small sample sizes and focused solely on specific neurological symptoms, limiting the generalizability of their findings (34). Besides, they relied on *a priori* selection of brain regions, which limits the identification of alternative areas that may be recruited into a network (7), and on time-windowed estimates, which confine the investigation to slow changes in connectivity (8). Here, we aimed to overcome these limitations by using a state-of-the-art data-driven dynamic method, the iCAP framework, which uses single frames to reveal large-scale brain networks that can be spatially and temporally overlapping. This method proved to be successful in disentangling the spatiotemporal organization of brain (23) and spinal cord activity (26) in able-bodied subjects. Deploying it in an extensive cohort of stroke patients (n = 103) with heterogeneous neurological syndromes and scanned longitudinally, we uncovered a temporal imbalance of network recruitment compared to healthy controls and showed how these dynamic changes articulate with anatomical and behavioral disruptions.

### Delving into network dynamics

We deployed the iCAP framework and found sixteen large-scale brain networks, or iCAPs, whose spatial patterns were in agreement with previous literature (23–26). This spatial organization was robust in healthy subjects and in stroke patients, with similar patterns in the two populations. In particular, networks were bilateral regardless of the side of the lesion, suggesting a preserved organization across brain regions. While this might seem in contradiction with the decrease in interhemispheric connectivity previously reported post-stroke (18, 35–37), it should be noted that fine-grained coordination between brain areas can be occulted when using classical *static* FC approaches (13, 25). For instance, the correlation between distinct brain areas exhibiting both positive and negative correlations in distinct time-windows would result in a low *static* functional connectivity, in spite of the dynamic synchronization existing between these regions (25). Therefore, it follows that a decrease in FC does not necessarily imply a decrease in global network activity. However, decreased FC might reflect an overall reduction in the efficiency of information transfer, which could manifest in the form of aberrant durations of functional networks (12, 25, 31).

To evaluate these time-varying properties, we thus derived subject-specific iCAP time-courses and computed average durations of activation for each individual iCAP, a measure that cannot be explained in terms of *static* connectivity. Of note, a similar approach previously allowed to show the link between brain dynamics and risk factors for schizophrenia in 22q11.2 deletion syndrome (24). In our study, network durations were stable over time in healthy subjects, while durations were, in contrast, affected post-stroke. Specifically, patients exhibited a varied pattern of longer and shorter network activations alluding to an “under-engagement” in certain brain states coupled with an “over-engagement” in others. These results, combined with the preserved overall amount of dynamic fluctuations (i.e., similar number of significant innovation frames between patients and healthy subjects), hint at a dynamical imbalance following stroke, rather than an overall shift towards a slower or a faster brain activity. A compelling observation was that networks usually associated in static analyses, such as the two sensorimotor networks (iCAPs 3 and 8) presented opposite engaging behaviors after lesions. This suggests that intricate and non-stationary behavior are also present within overlapping brain regions, something that would be overlooked when relying on conventional *static* methods.

### Structural substrates of disrupted dynamics

We then assessed whether this post-stroke imbalance was determined by structural damages. Indeed, understanding how the human brain orchestrates large-scale functional networks despite being constrained by a rigid anatomical substrate and how this orchestration is affected by lesions is still an open question in clinical neuroscience (38–41). For this, we related iCAP temporal properties to structural measures of disconnections (27, 37) and found that altered durations were explained by the disruption of white matter tracts, particularly within the projection pathways, as well as within the cingulum and the commissural pathways. The former subserve the subcortical integration of sensorimotor inputs and outputs between the cerebral cortex and the peripheral system. The latter, instead, connect cerebral hemispheres. Both these pathways are involved in providing local and global efficiency (42), measures that are indicative of the capacity for parallel information transfer and integrated processing between, respectively, short and long distance regions (43). As such, disruptions of these tracts likely drive the widespread neurobiological processes occurring in the course of stroke recovery (31).

### Domain-specific behavioral correlates

Finally, we assessed whether these lesion-induced changes in network dynamics correlate with behavioral impairments and recovery. To this end, we capitalized on the longitudinal nature of the dataset so as to assess short- and long-term recovery processes and on its multi-domain clinical assessments. Earlier observations indicated that most deficits restore within three months, with additional recovery, yet more limited, up to one year following lesion (30). Interestingly, these multi-scale recovery patterns were reflected in the temporal dynamics of brain activity, as the initial imbalance observed in severely affected patients normalized towards the level of healthy subjects, mostly within the first three months. Thanks to the heterogeneous patient population, we showed that these trends were significant only for attention, language and spatial memory disorders, as also observed for post-stroke decrease in network modularity in an earlier study (31). This suggests that both modularity and network duration are associated with higher cognitive deficits, in the form of functional changes likely reflecting inter-hemispheric homotopic integration and within-hemisphere segregation (18). It is noteworthy that these alterations in static connectivity were not reflected in the spatial organization of the large-scale networks but on their temporal durations. Motor deficits, instead, are known to be better predicted by lesion location (18), probably indicating that these functions rely less on information integration between large-scale networks. Further supporting the differential nature of impairments, changes in duration were network-specific (e.g., only durations of auditory and visual networks correlated with language deficits). Finally, deficits and recovery were associated with distant brain regions, bringing new evidence to corroborate that stroke is not a focal disease but, instead, a network disease (20, 22).

### Clinical considerations

In summary, we found that durations of large-scale resting-state networks was altered in consequence of interruptions in white-matter tracts. The brain was, therefore, less resilient and modular after stroke, as previously hypothesized (31). Importantly, the restoration of the networks’ natural temporal properties, likely accompanied by a restoration of this resilience, was crucial to achieve behavioral recovery, especially for cognitive deficits. Altogether, these results underscore the clinical relevance of network temporal properties, in particular as regards their duration. Therefore, dynamic functional connectivity methods seem crucial not only to elucidate pathophysiology changes post-stroke but also to capture potential therapeutic targets that could possibly be modulated, for instance using non-invasive brain stimulation. While such interventions have been frequently employed to improve motor recovery after stroke (22, 44, 45), only a restricted number of attempts have explored brain stimulation to treat post-stroke cognitive deficits (46), despite several successful applications in psychiatry (47). In this field, recent investigations jointly considering anatomy and functional dynamics provided valuable insights to guide neuromodulation treatments (48, 49). In this regard, we foresee that investigating stroke-related disruptions using dynamic approaches could help shed light on meaningful temporal properties, which are directly tied to the underlying anatomical substrate, and that can be more readily leveraged to tune neuromodulation parameters. Considering that brain stimulation approaches can change excitability of functional connections within and between cortical areas with a high temporal and topographical resolution (22), tuning parameters based on temporal properties could enable a better modulation of large-scale brain networks with the aim to improve patient’s clinical outcome. In this context, the advent of multimodal recording and stimulation techniques such as TMS-fMRI is critical to first probe the effects of brain modulation over network dynamics (21, 45, 50), so that stimulation paradigms can be tuned accordingly. Finally, it is noteworthy that dFC methods do not require additional acquisition constraints for the patients. As such, clinical transitions to fMRI time-resolved methods appears not only necessary but also technologically possible.

### Materials and Methods

#### Participant information

All participants gave their written informed consent to participate in accordance with the Declaration of Helsinki, and the study was approved by the Institutional Review Board at Washington University in St. Louis. The complete data collection protocol is described in detail elsewhere (29). Data from 127 first-time stroke patients with clinical evidence of impairment and data from 21 demographically matched healthy controls were considered for inclusion in the analyses (**Table S1**). Healthy adults matched the stroke population by age, gender, handedness, and level of education (see (18) for details in inclusion and exclusion criteria for the two population groups).

### Experimental protocol

Patients underwent a maximum of three testing sessions (**Table S1**): within 1-2 weeks (n = 103), 3 months (n = 72), and 12 months (n = 54) after the lesion (see (31) for motivations and percentages of patients lost over the three time points). Healthy control subjects participated in two testing sessions three months apart. Each testing session consisted of seven resting state fMRI runs, each including 128 volumes (30 min total) and a neuropsychological assessment. Imaging and behavioral testing sessions were usually performed on the same day.

### Neuropsychological Assessment

Participants underwent a comprehensive battery of 44 behavioral tests across four behavioral domains language, memory, motor, attention and visual function, chosen to represent a wide range of the most commonly identified deficits in stroke patients. Scores were only recorded for tasks that subjects were able to complete. Therefore, different domains include different numbers of subjects (**Table S1**). In order to isolate clusters of deficits based on these different measures, we applied principal component analysis (PCA) on the behavioral data for each domain separately (e.g., motor, language, attention, spatial memory, and verbal memory) following our previous method (29, 30) (see **SI** for details).

### Imaging acquisitions

All imaging was performed using a Siemens 3T Tim-Trio scanner and a standard 12-channel head coil. The MRI protocol included structural and functional scans. Structural scans included: (i) a sagittal T1-weighted MP-RAGE (TR = 1950 msec, TE = 2.26 msec, flip angle = 90°, voxel size = 1.0 × 1.0 × 1.0 mm); (ii) a transverse T2-weighted turbo spin echo (TR = 2500 msec, TE = 435 msec, voxel size = 1.0 × 1.0 × 1.0mm); and (iii) sagittal fluid attenuated inversion recovery (FLAIR) (TR = 750 msec, TE = 32 msec, voxel size = 1.5 × 1.5 × 1.5 mm). Resting state functional scans were acquired with a gradient echo EPI sequence (TR = 2,000 msec, TE = 2 msec, 32 contiguous 4-mm slices, 4 × 4 mm in-plane resolution) during which participants were instructed to fixate a small cross white cross centered on a screen with a black background in a low luminance environment.

### fMRI data pre-processing

MRI scans were pre-processed using a pipeline adapted from our previous (23–25) studies that used SPM8 (http://www.fil.ion.ucl.ac.uk/spm/). After realignment of functional scans, we applied spatial smoothing with an isotropic Gaussian kernel of 5-mm full-width at half-maximum and coregistered structural scans to the functional mean. Individual tissue (white and gray matter, and cerebrospinal fluid (CSF)) maps were segmented from the T1 image. Then, the first ten functional scans were discarded, resulting in T = 118 volumes per run (for a total of 826 volumes per subject – i.e., 118 × 7 runs). The following sources of spurious variance were regressed out from these BOLD time series: (i) six parameters obtained by rigid body correction of head motion, (ii) average white matter and CSF signals. Finally, for each session fMRI runs were spatially realigned to the functional mean of the first run.

### Extraction of large-scale brain networks

In order to extract large-scale brain networks and their temporal characteristics we deployed the iCAP pipeline (23) (**Figure S2** and **SI**). We first applied the Total Activation framework (TA, (53)), which applies hemodynamically informed deconvolution to the pre-processed fMRI time series (native space) of each run separately to reliably retrieve activity-inducing signals. Then, for each subject and session, activity-inducing time courses of all runs were concatenated, removing three volumes at each intersect (i.e., 808 volume kept in total per subject and session) and significant activation change-points (i.e., transients) were computed as the temporal derivative of these activity-inducing signals. Transients were then normalized to the MNI space considering the lesion mask as a prior for an additional tissue class in the segmentation procedure (54), concatenated across all subjects (i.e., patients and healthy subjects – see **SI** for subjects excluded from the analysis) and sessions, and fed into a temporal k-means clustering to obtain large-scale resting-state networks, the iCAPs. The optimum number of 16 clusters was determined by evidence accumulation (55, 56) (**Figure S3a** and **SI**). Finally, iCAP time courses were obtained for each subject and session by transient-informed spatiotemporal back-projection of the 16 spatial maps onto the activity-inducing signals (**Figure S3b** and (57)).

### Extraction of Temporal Properties

iCAP time courses were Z-scored within each subject and session and thresholded at a Z score > |1| to determine “active” time points. The choice of threshold was motivated by previous works that implemented TA and iCAP framework (23, 57). For each iCAP, we computed the total duration of overall activation as percentage of the total scanning time within a session.

### White matter tracts disconnections

In order to quantify the damages to the different cortical and subcortical white matter tracts, we used our previous approach (27, 37). Specifically, for each patient, we intersected the lesion mask (see **SI**) with a streamline tractography atlas and we quantified the proportion of streamlines disconnected for each tract. For the tractography atlas, we used a publicly available diffusion MRI streamline tractography atlas (42), which consisted of 70 tracts: 65 neuroanatomically defined fiber bundles corresponding to commissural, association, projection, brainstem, and cerebellar pathways (cranial nerves were not included), and the corpus callosum split into 5 segments (27, 37).

### Quantification and statistical analysis

#### Stability of iCAP spatial patterns

First, we obtained iCAP spatial patterns for each individual and session by averaging significant innovation frames belonging to the corresponding subject and session. For each iCAP, we then computed iCAP stability as the average mean cosine similarity between the cluster centroid (i.e., the global iCAP map) and the individual subject iCAP maps.

#### Group comparisons of iCAP spatial patterns

For each group and testing sessions, iCAP spatial patterns were obtained by averaging significant innovation frames belonging to the corresponding group and session. The group- and/or session-specific spatial patterns were then compared using cosine similarity.

#### Group comparisons of iCAP durations

In order to compare iCAP durations between patients and healthy control subjects, we deployed a Bayesian classifier, specifically a LDA (33). We built a two-class LDA classifier (accounting for different covariance matrices for each class) using the durations of the 16 iCAPs. To rank the features of the classifier, we calculated the discrimination power for the two classes (i.e., patients and healthy controls) for each feature separately, using a two-sample Mann-Whitney test. We performed leave-one-subject-out cross-validation, leaving out, for each fold, all the sessions of one subject (either healthy control or patient) and we ranked the features by their absolute standardized u-statistic obtained from the Mann-Whitney test. We selected the features that were stable over folds (i.e., the features that were always ranked as most discriminative, n = 6) and used them to build an LDA classifier. Decoding accuracy values were averaged over leave-one-out cross-validation folds. Receiver operating characteristic (ROC) curves and confusion matrices were computed considering all folds and the area under the ROC curve (AUC) was computed. To assess the statistical significance of the classification, we built N=1000 classifiers with randomly assigned labels at each permutation and estimated AUC and classification accuracy for each permutation. For both measures, p-values were obtained as 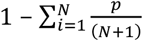 where *p* corresponds to the cases where the original AUC and/or classification accuracy are above random AUC and/or classification accuracy.

#### Partial Least Squares Correlation

We deployed partial least squares correlation (PLSC) (24, 58–60) to evaluate multivariate patterns of correlation between iCAP durations and: (i) white matter disconnections; (ii) behavioral scores and recovery (**SI** for details).

i. we conducted a PLSC analysis with duration of the 16 iCAPs as brain variables and with white matter tract disconnections as anatomical variables. Microstructural changes in the weeks/months following the stroke can be reasonably expected because of inflammatory reactions as well as degenerative and neuroplastic processes, leading to further changes in the anatomical disconnections. However, white matter disconnections were obtained only from structural images obtained at 1-2 weeks post-lesion. Therefore, for the PLSC we considered brain and anatomical data only for this first time point. Our assumption was further justified by non-significant correlation components for later time points.
ii. we computed a group PLSC analysis between the 16 iCAP durations and the behavioral scores for three groups: patients without severe acute deficits, patients with severe acute deficits, and healthy controls as normative data. PLSC analysis was computed for each neuropsychological domain (attention, spatial memory, verbal memory, language, and motor) separately, as only a limited number of participants performed the assessment for all four classes (**Table S1**). Behavioral scores were obtained from the first PCA component. In order to capture multivariate patterns of correlation with the behavioral measures and the recovery, for each subject and each time point the behavioral design matrix included *(i)* a variable containing the average value of the behavioral score over all subject’s time points (mean), *(ii)* a variable containing the difference between the behavioral score at that time point and the average value over all time points (delta), and (iii) a variable for the mean by delta interaction.

## Data Availability

The full set of neuroimaging data (along with behavioral data) are available at http://cnda.wustl.edu/app/template/Login and the scripts for the iCAP framework and PLSC analysis are openly accessible (https://c4science.ch/source/iCAPs/ and https://github.com/danizoeller/myPLS, respectively). Other specific data and scripts are available upon request to the authors.

## Acknowledgments

The authors would like to thank Drs. Anjali Tarun and Daniela Zöller for the inspiring discussions and Drs. Giulia Preti and Raphaël Liégeois for their comments on the manuscript.

## Notes

### Competing Interest Statement

The authors have declared no competing interest.

### Clinical Trial

This study was not a clinical trial, but an observational study.

### Funding Statement

No external funding was received for this study

### Author Declarations

All participants gave their written informed consent to participate in accordance with the Declaration of Helsinki, and the study was approved by the Institutional Review Board at Washington University in St. Louis.

