## Supplementary material for "Post-stroke reorganization of transient brain activity characterizes deficits and recovery of cognitive functions"

### Supplementary Material and Methods

### Participants exclusion criteria

From the initial sample of 148 subjects, N = 24 patients and n = 2 healthy subjects were excluded from the analysis because of excessive motion (> 60% of volumes with a framewise displacement of more than 0.5 mm (1)) or because of a low number of significant innovation frames (< 131 significant innovation frames over a total of 808 volumes).

### Behavioral domain scores

Dimensionality reduction was performed on the performance data as described in detail in (2, 3). First, tasks were categorized as attention, spatial memory, verbal memory, language, and motor. A PCA was run on the behavioral data of the session at 1-2 weeks post-lesion and the first component was used as a domain score for each category separately (see **SI** for a description of the components). The component scores for subsequent time points and for the age-matched controls were generated by normalizing the original data based on the sub-acute values and projecting them in the PCA space. Then, each of the components and time-points was z-scored based on the first measurement of the healthy control group, allowing comparisons across timepoints and behavioral domains. Patients with a score >/< 2 standard deviations were identified as "patients with/without severe acute deficits". For each behavioral domain, we followed the same procedure as (3) and conducted an ANOVA across the three timepoints comparing patients with and without severe acute deficits. In attention, the first component described 24.7% of variance and was strongly related to measures of attentional field bias such as the total number of miss items in Mesulam cancellation test (r = 0.61), and the accuracy (r = 0.85) and reaction time (r = 0.72) in the Posner task. For the motor domain, the first two components (explained variance 40.0%

and 32.2% respectively) correlated with left and right motor function. In language the first component accounted for 77.3% of the variance and correlated with tasks of auditory comprehension, expression, and reading (r > 0.81). Finally, for spatial memory the first component (explained variance 61%) was correlated with measures of visuospatial memory such as immediate (r = 0.87) and delayed recall (r = 0.90) of visual information on the Brief Visuospatial Memory Test; whereas for verbal memory the first factor (explained variance 74.9%) correlates with measures from the Hopkins Verbal Learning Test such as delayed recall of words (r = 0.96).

### Lesion masking

Individual T1 MRI images were registered to the MNI brain using FSL (FMRIB Software Library) FNIRT tool (4). Lesions were manually segmented on individual structural MRI images obtained 1–2 weeks post-lesion using the Analyze biomedical imaging software system (5). Special attention was given to distinguish lesion from CSF, hemorrhage from surrounding vasogenic edema, and to identify the degree of periventricular white matter damage present. In hemorrhagic strokes, edema was included in the lesion. The staff that was involved in segmenting or in reviewing the lesions (M.C. and Alexandre Carter) was blind to the individual behavioral data.

### Total Activation and iCAPs

The Total Activation (TA) and iCAPs framework is based on the detection of significant changepoints in deconvolved fMRI time series. Matlab code for the application of the whole framework can be found at https://www.c4science.ch/source/iCAPs.
TA was applied for each subject and session to each run separately in order to obtain activity-inducing time courses. For each subject, activity-inducing time courses were contacted over all runs and activation change-points were computed as the temporal derivative of these activity-inducing signals. In order to select significant innovations frames (i.e., frames with significant transitioning activities - transients), a two-step thresholding procedure was employed with temporal and spatial thresholds selected based on previous work (6–8): i) temporal thresholding - for each voxel, a surrogate distribution was obtained by applying TA on phase randomized data and a 1% confidence interval was used to select significant voxels; ii) spatial thresholding - only the innovation frames with at least 5% of active voxels were considered to be significant. Transients were then normalized to Montreal Neurological Institute (MNI) and concatenated across all subjects (i.e., patients and healthy subjects) and sessions, and fed into a temporal k-means clustering to obtain large-scale resting-state networks, the iCAPs. The optimum number of clusters was determined by evidence accumulation (9, 10) (see **Figures S3a**). Briefly, the k-means clustering was done for K values ranging from 10 to 24, in order to obtain a co-association matrix summarizing how often two frames were clustered together. A second phase of clustering was performed using this co-association matrix and, this time, hierarchical clustering with two different linkage functions (average and weighted). We then computed the percentage of agreement between these two linkage functions, as well as with the k-means solution. The number of iCAPs was chosen as the number that showed the highest percentage of agreement, thus resulting in the extraction 16 iCAPs.

### Partial Least Squares Correlation

Partial least squares correlation (PLSC) has been previously successfully employed to characterize covarying patterns of structural and functional connectivity in healthy individuals (11), and it is nowadays considered a clinically relevant method (7, 12–15). It seeks to define linear combinations of two data matrices (**X**, i.e., the brain networks properties – iCAP durations, and **Y,** i.e., the behavioral or anatomical variables) that maximally explain the covariance between the two matrices. The first step in PLSC is the computation of the correlation matrix between **X** and **Y** (**R = X'Y**). In our approach, **X** and **Y** were z-scored across subjects before correlation. Then **R** is decomposed in *N* latent variables, or "correlation components" (where *N* is the minimum number between the number of included behavioral/anatomical variables and the number of iCAPs), using singular value composition **R=USV'** with **U'U=V'V=I**. Each correlation component has a singular value (on the diagonal of **S**) that specifies the explained correlation, as well as *Nx* iCAP durations saliences or "duration weights" (rows of **V'**) and *Ny* behavioral/anatomical saliences or “behavioral/anatomical weights” (columns of **U**). The saliences (which lie between -1 and 1) indicate how strongly each variable contributes to the multivariate behavioral-brain/anatomical-brain correlation in a certain correlation component. We can then compute the so called “brain scores” by projecting every individual’s iCAP durations onto the respective brain weights with **Lx=XV**. We used permutation testing with 1000 permutations to evaluate if any of the correlation components was significant and bootstrapping with 500 bootstrap samples with replacement to evaluate the stability of the behavior/anatomical and brain weights. Brain and behavioral saliences were recalculated for every bootstrap sample, resulting in a typical bootstrap distributing of the salience values. Saliences were considered significant if lower/higher than lower/upper bound of 95% confidence interval of bootstrapping distributions.

Group PLSC analysis, as the one used for the correlation between iCAP durations and behavioral scores, entails that a correlation matrix is computed per group (in this case healthy controls $\mathbf{R_{HC}}$, patients with less severe acute deficits $\mathbf{R_{ND}}$, and patients with severe acute deficits $\mathbf{R_{D}}$). The common correlation matrix R is then computed by concatenating $\mathbf{R_{HC}}$, $\mathbf{R_{ND}}$ and $\mathbf{R_{D}}$ resulting in $3Ny$ behavior saliences.

### Data availability

As the clinical effectiveness of all these methods is critically dependent on the availability of publicly released tools (16), the full set of neuroimaging data (along with behavioral data) are available at http://cnda.wustl.edu /app/template/Login and the scripts for the iCAP framework and PLSC analysis are openly accessible (https://c4science.ch/source/iCAPs/ and https://github.com/danizoeller/myPLS, respectively). Other specific data and scripts are available upon request to the authors.

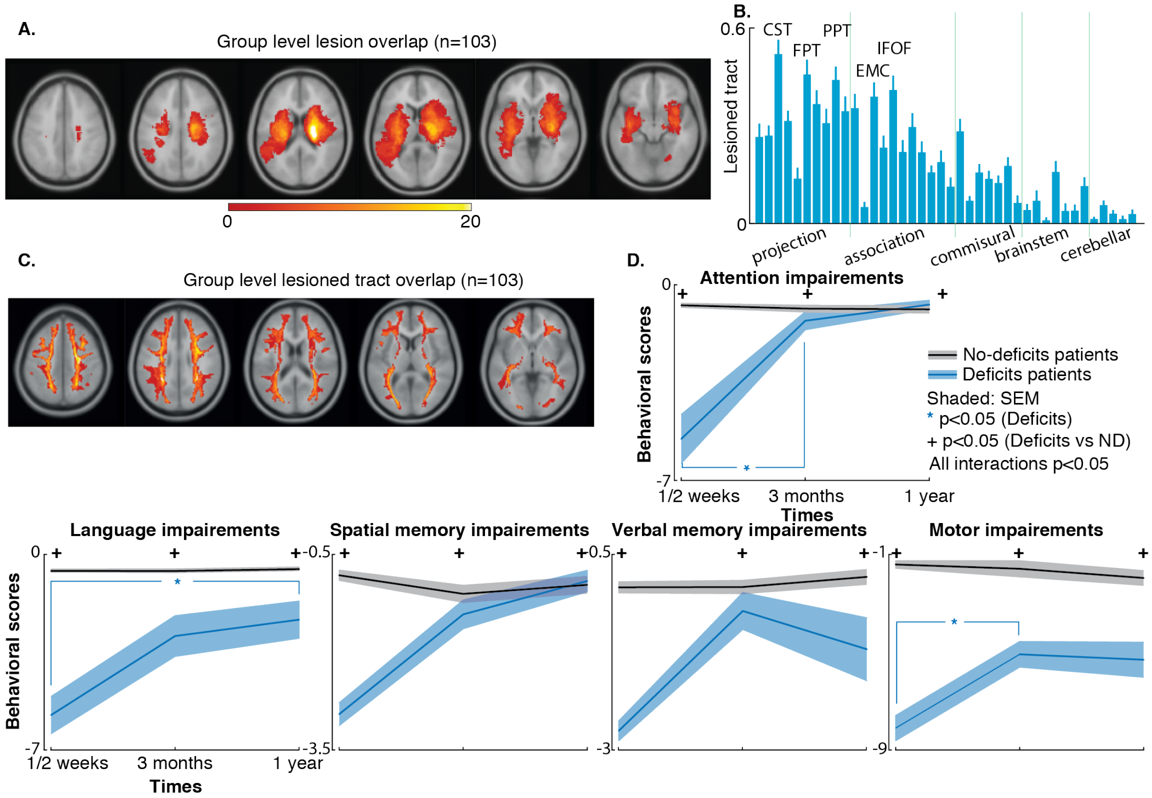


**Fig. S1. Patient population | A.** Topography of stroke. Binary lesion masks of all 103 stroke patients included in the analysis were summed and overlaid in Montreal Neurological Institute coordinates. **B.** Mean ± SEM over subjects of the portion of streamlines disconnected for each tract. Tracts were grouped in projection pathways, association pathways, commissural pathways, brain- stem pathways, and cerebellar pathways following (17). **C.** Streamlines disconnected overlay map in Montreal Neurological Institute coordinates for the 103 stroke patients included in the analysis. **D.** Time course of recovery for each domain separately (e.g., attention (n=81 subjects), language (n=96), spatial memory (n=76), verbal memory (n=76), and motor (n=89)). We reported behavioral factor scores over the three time points for patients that have severe deficits (blue lines) and did not (black lines) have severe deficit 1-2 weeks post stroke (mean ± SEM over subjects). Interactions between the two groups over time are significant for each domain (as indicated by + symbols). * represents significant differences (corrected for multiple comparisons) within timepoints for patients that did have severe acute deficits.

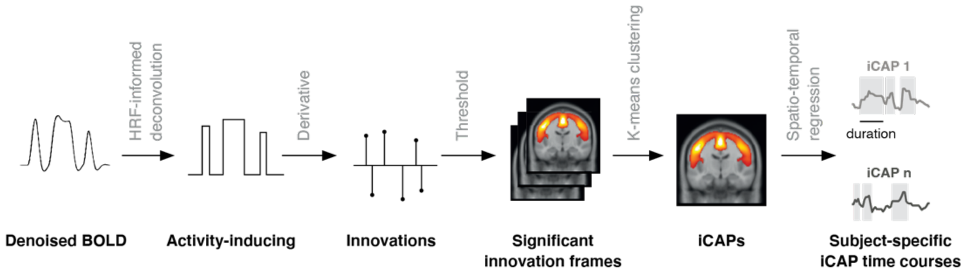


**Fig. S2. Dynamic functional connectivity framework |** Functional images from individual subjects are denoised to circumvent the effect of various sources of noise. The hemodynamic blur is subsequently removed using hemodynamic-informed deconvolution, which reveals the activity-inducing signals. The innovation frames are then obtained by temporal derivation. A two-step thresholding (temporal and spatial) is applied to select significant innovation frames (i.e., transients), which undergo temporal clustering over subjects to obtain stable iCAPs. The latter are fitted back to the individual activity inducing signals to recover temporal profiles of the iCAPs for further time-resolved analysis. For each participant, session, and iCAP, we then computed the average duration over the total acquisition length.

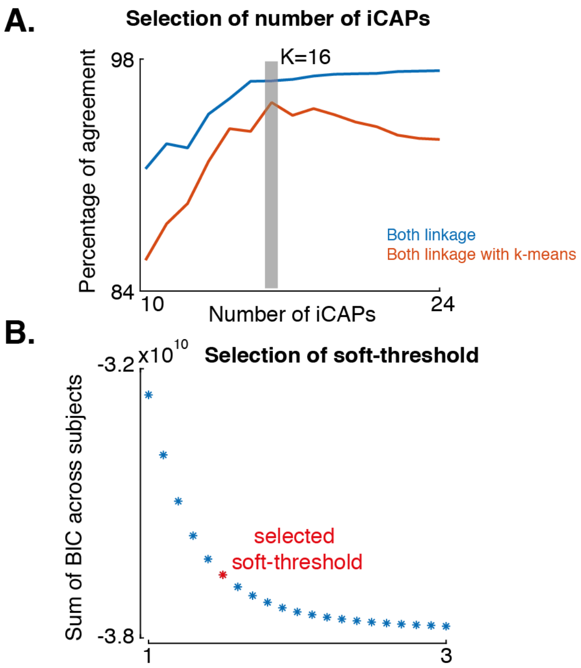


**Fig. S3. Parameters selections for the dynamic functional connectivity framework | A.** Percentage of agreement for linkage (blue) and linkage with k-means (orange) for number of iCAPs from 10 to 24. The number of iCAPs was chosen as the number at which both methods had the highest percentage (i.e., k=16 iCAPs). **B.** Evaluation of soft assignment factors $\xi$ from 1 (hard cluster assignment) to 3 (all iCAPs allowed to change at timepoints of significant transients). The red dot indicates the selected soft-threshold (i.e.,$\boldsymbol{\xi} = \mathbf{1.5}$) (see (18) for details).

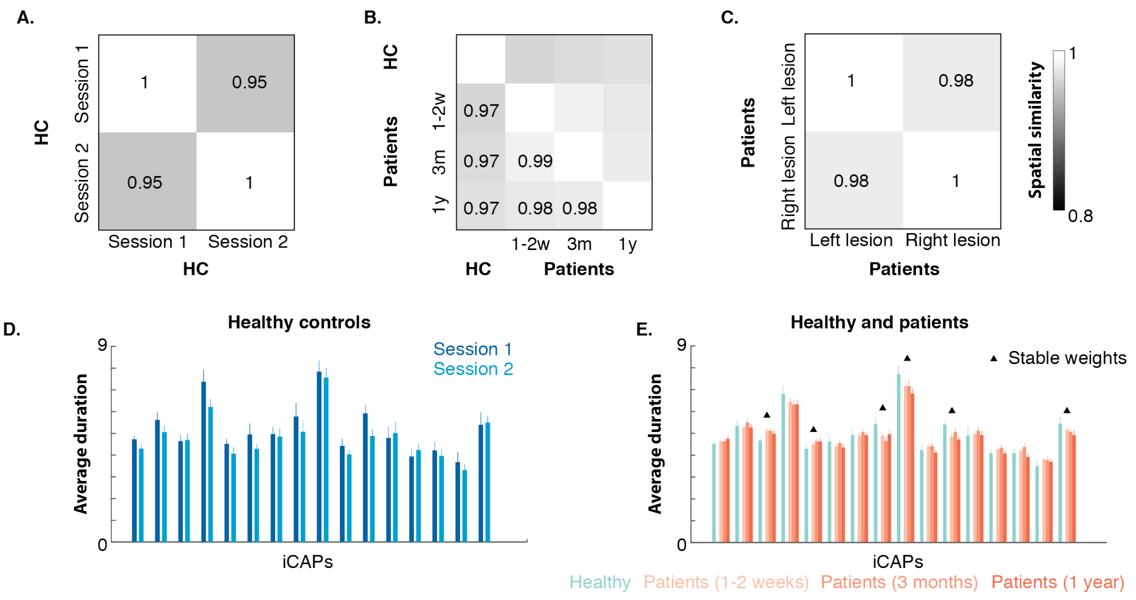


**Fig. S4. Stability of spatial patterns and average duration | A.** Cosine similarity between iCAPs spatial patterns of healthy control subject's session 1 and session 2 averaged over the 16 iCAPs. **B.** Cosine similarity between iCAPs spatial patterns for healthy control subjects and stroke patients at 1-2 weeks, 3 months, and one-year post-lesion averaged over the 16 iCAPs. **C.** Cosine similarity between iCAPs spatial patterns of patients with lesion in the left hemisphere and patients with lesion in the right hemisphere. **D.** iCAPs average duration for healthy control subject's session 1 (dark blue) and session 2 (light blue) (mean ± SEM over subjects). **E.** iCAPs average duration for healthy control subjects averaged over session 1 and 2 (cyan), patients at 1-2 weeks (light orange), 3 months (orange), and one-year post-lesion (dark orange) (mean ± SEM over subjects). Triangle indicates iCAPs that have stable weights over folds of the LDA classifier.

**Table S1. Patients demographics |** Age, Sex, Dominant Hand, Education, Lesion Side, Lesion Type, number of participants included for the 5 different behavioral domains (Attention, Spatial Memory, Verbal Memory, Motor, and Language), and time between lesion onset and scans at different time points $T_0$ (1/2 weeks), $T_1$ (3 months), and $T_2$ (1-year post-lesion). SD: Standard Deviation; M: Male; F: Female; R: Right; L: Left; I: Ischemic; H: Hemorrhagic; O: other stroke etiologies (e.g. hemorrhagic conversion, vertebral artery dissection, etc.); Att: Attention; Mem: Memory; Mot: Motor; Lan: Language.

| Group | Age (Mean/SD) | Sex | Hand | Education (Mean/SD) | Lesion Side | Lesion Type | Att. | Spatial Mem. | Verbal Mem. | Mot. | Lang. | Time (Mean/SD) |
|---|---|---|---|---|---|---|---|---|---|---|---|---|
| **Patients (N=103)** | 53.5/10.2 | 59M/ 44F | 95R/8L | 13/2.5 | 47R/56L | 80%I<br>15%H<br>5% O | 81 | 76 | 76 | 89 | 96 | $T_0$: 13.4d/4.8d<br>$T_1$: 112.5d/8.4d<br>$T_2$: 395.5d/55.1d |
| **Controls (N=19)** | 52.7/12.8 | 7M/ 12F | 17R/2L | 13.8/2.4 | N/A | N/A | 19 | 19 | 19 | 19 | 19 | N/A |

**Table S2. iCAPs |** iCAP functional networks of Greicius atlas (19), iCAP regions in the automated anatomical labeling (AAL) atlas (20) and 34 regions corresponding to portions of the cerebellum, thalamus, and basal ganglia taken from the automatic anatomical labeling (AAL) atlas. Percentiles indicate the fraction of voxels of a functional network or region that have a z-score > 1.9. A network/region is listed if more than 20% of the network/region is included in the iCAP.

| iCAP | Greicius network (%) | AAL Lobe | AAL Region (%) | z-score | N vox |
|---|---|---|---|---|---|
| **1** | Primary visual (99%) | Occipital | Cuneus_L (67%) | 3.19 | 298 |
| | Precuneus (63%) | Occipital | Cuneus_R (65%) | 3.16 | 283 |
| | DMN (33%) | Occipital | Calcarine_L (53%) | 3.31 | 343 |
| | | Occipital | Calcarine_R (63%) | 3.26 | 340 |
| | | Occipital | Lingual_L (32%) | 2.79 | 211 |
| | | Occipital | Lingual_R (28%) | 2.80 | 188 |
| | | Occipital | Occipital_Sup_L (23%) | 2.53 | 91 |
| | | Parietal | Precuneus_L (30%) | 2.73 | 326 |
| | | Parietal | Precuneus_R (35%) | 2.83 | 324 |
| **2** | High visual (46%) | Occipital | Occipital_Inf_L (30%) | 2.46 | 80 |
| | Cerebellum(26%) | Occipital | Occipital_Inf_R (25%) | 2.51 | 77 |
| | | Occipital | Fusiform_L (23%) | 2.74 | 153 |
| | | Occipital | Lingual_R (21%) | 2.72 | 142 |
| | | Cereb | Cerebellum_Crus1_L (50%) | 2.58 | 380 |
| | | Cereb | Cerebellum_Crus1_R (41%) | 2.41 | 327 |
| | | Cereb | Cerebellum_6_L (79%) | 2.56 | 407 |
| | | Cereb | Cerebellum_6_R (65%) | 2.44 | 351 |
| | | Cereb | Cerebellum_Crus2_L (30%) | 2.35 | 172 |
| | | Vermis | Vermis_6 (85%) | 2.39 | 89 |
| | | Vermis | Vermis_7 (95%) | 2.70 | 56 |
| | | Vermis | Vermis_8 (70%) | 2.33 | 47 |
| **3** | Sensorimotor (37%) | Parietal | Postcentral_L (44%) | 2.89 | 512 |
| | Auditory (27%) | Parietal | Postcentral_R (26%) | 2.60 | 297 |
| | Visuospatial (28%) | Parietal | Paracentral_Lobule_L (36%) | 2.76 | 154 |
| | | Parietal | Paracentral_Lobule_R (32%) | 2.61 | 72 |
| | | Limbic | Cingulum_Mid_L (38%) | 2.86 | 236 |
| | | Limbic | Cingulum_Mid_R (30%) | 2.87 | 180 |
| | | Frontal | Supp_Motor_Area_L (33%) | 3.06 | 214 |
| | | Frontal | Supp_Motor_Area_R (31%) | 2.98 | 207 |
| | | Frontal | Precentral_L (27%) | 2.67 | 277 |
| | | Frontal | Precentral_R (21%) | 2.65 | 202 |
| **4** | DMN (41%) | Frontal | Frontal_Sup_Medial_L (45%) | 2.97 | 383 |
| | Precuneus (22%) | Frontal | Frontal_Sup_Medial_R (39%) | 2.80 | 253 |
| | ECN (33%) | Frontal | Frontal_Sup_L (24%) | 2.80 | 254 |
| | Anterior Salience (45%) | Frontal | Frontal_Mid_L (29%) | 2.72 | 419 |
| | | Limbic | Cingulum_Ant_L (27%) | 2.69 | 117 |
| | | Limbic | Cingulum_Ant_R (36%) | 2.72 | 141 |
| | | Parietal | Angular_L (27%) | 2.51 | 92 |
| **5** | Auditory (93%) | Temporal | Heschl_L (94%) | 3.50 | 68 |
| | Language (22%) | Temporal | Heschl_R (92%) | 3.75 | 67 |
| | | Temporal | Temporal_Sup_L (57%) | 3.05 | 386 |
| | | Temporal | Temporal_Sup_R (54%) | 3.03 | 521 |
| | | Central | Rolandic_Oper_L (67%) | 3.16 | 203 |
| | | Central | Rolandic_Oper_R (70%) | 3.16 | 281 |
| | | Limbic | Insula_L (68%) | 3.15 | 385 |
| | | Limbic | Insula_R (70%) | 3.27 | 375 |
| | | Frontal | Frontal_Inf_Oper_R (21%) | 2.76 | 90 |
| **6** | Primary visual (99%) | Occipital | Calcarine_L (52%) | 2.85 | 336 |
| | High visual (64%) | Occipital | Calcarine_R (68%) | 2.92 | 368 |
| | | Occipital | Lingual_L (65%) | 2.94 | 435 |

| | | | | | |
|---|---|---|---|---|---|
| | | Occipital | Lingual_R (68%) | 3.00 | 463 |
| | | Occipital | Occipital_Mid_L (22%) | 2.59 | 211 |
| | | Occipital | Occipital_Mid_R (22%) | 2.72 | 133 |
| | | Occipital | Occipital_Inf_L (30%) | 2.64 | 81 |
| | | Occipital | Occipital_Inf_R (21%) | 2.67 | 66 |
| | | Occipital | Fusiform_R (25%) | 2.76 | 188 |
| | | Cereb | Cerebellum_4_5_L (30%) | 2.30 | 106 |
| | | Cereb | Cerebellum_4_5_R (31%) | 2.30 | 76 |
| | | Cereb | Cerebellum_6_L (59%) | 2.51 | 306 |
| | | Cereb | Cerebellum_6_R (57%) | 2.43 | 304 |
| | | Vermis | Vermis_4_5 (63%) | 2.41 | 116 |
| | | Vermis | Vermis_6 (96%) | 2.64 | 101 |
| | | Vermis | Vermis_7 (63%) | 2.07 | 37 |
| **7** | Cerebellum (22%) | Limbic | Hippocampus_L (52%) | 2.20 | 143 |
| | Amygdala (23%) | Limbic | Hippocampus_R (65%) | 2.29 | 188 |
| | | Limbic | ParaHippocampal_L (27%) | 2.46 | 77 |
| | | Limbic | ParaHippocampal_R (30%) | 2.62 | 94 |
| | | Occipital | Lingual_L (26%) | 2.25 | 172 |
| | | Occipital | Lingual_R (29%) | 2.29 | 198 |
| | | Occipital | Fusiform_L (27%) | 2.30 | 184 |
| | | Occipital | Fusiform_R (43%) | 2.37 | 328 |
| | | Cereb | Cerebellum_3_L (35%) | 2.23 | 14 |
| | | Cereb | Cerebellum_3_R (43%) | 2.39 | 27 |
| | | Cereb | Cerebellum_4_5_L (71%) | 2.62 | 256 |
| | | Cereb | Cerebellum_4_5_R (86%) | 2.71 | 208 |
| | | Cereb | Cerebellum_6_L (71%) | 2.33 | 367 |
| | | Cereb | Cerebellum_6_R (72%) | 2.43 | 389 |
| | | Vermis | Vermis_3 (40%) | 2.25 | 27 |
| | | Vermis | Vermis_4_5 (75%) | 2.57 | 138 |
| | | Vermis | Vermis_6 (88%) | 2.47 | 92 |
| | | Vermis | Vermis_7 (51%) | 2.14 | 30 |
| | | Vermis | Vermis_8 (37%) | 2.04 | 25 |
| | | Vermis | Vermis_9 (40%) | 2.00 | 23 |
| **8** | Sensorimotor (33%) | Frontal | Precentral_L (30%) | 2.83 | 308 |
| | Ventral DMN (26%) | Frontal | Precentral_R (37%) | 3.00 | 370 |
| | Anterior Salience (24%) | Frontal | Supp_Motor_Area_L (68%) | 2.89 | 445 |
| | Visuospatial (27%) | Frontal | Supp_Motor_Area_R (76%) | 3.00 | 504 |
| | | Frontal | Frontal_Sup_R (21%) | 2.95 | 245 |
| | | Parietal | Postcentral_L (26%) | 2.74 | 299 |
| | | Parietal | Postcentral_R (37%) | 3.12 | 420 |
| | | Parietal | Parietal_Sup_L (25%) | 2.55 | 160 |
| | | Parietal | Parietal_Sup_R (32%) | 2.72 | 208 |
| | | Parietal | Precuneus_L (30%) | 2.72 | 318 |
| | | Parietal | Precuneus_R (23%) | 2.71 | 214 |
| | | Parietal | Paracentral_Lobule_L (68%) | 3.26 | 287 |
| | | Parietal | Paracentral_Lobule_R (70%) | 3.32 | 125 |
| | | Parietal | Parietal_Inf_R (30%) | 2.51 | 419 |
| | | Limbic | Cingulum_Mid_L (29%) | 2.42 | 182 |
| | | Limbic | Cingulum_Mid_R (24%) | 2.46 | 148 |
| **9** | Amygdala (78%) | Frontal | Frontal_Sup_Orb_L (26%) | 2.24 | 105 |
| | | Frontal | Frontal_Sup_Orb_R (42%) | 2.25 | 132 |
| | | Frontal | Frontal_Inf_Orb_L (41%) | 2.31 | 205 |
| | | Frontal | Frontal_Inf_Orb_R (31%) | 2.32 | 154 |
| | | Frontal | Olfactory_L (51%) | 2.21 | 44 |
| | | Frontal | Olfactory_R (53%) | 2.24 | 43 |
| | | Frontal | Rectus_L (88%) | 2.34 | 230 |
| | | Frontal | Rectus_R (91%) | 2.35 | 199 |
| | | Frontal | Frontal_Sup_Orb_Medial_L (21%) | 2.06 | 47 |
| | | Limbic | Hippocampus_L (42%) | 2.92 | 116 |

| | | | | | |
|---|---|---|---|---|---|
| | | Limbic | Hippocampus_R (46%) | 2.94 | 132 |
| | | Limbic | ParaHippocampal_L (35%) | 2.80 | 99 |
| | | Limbic | ParaHippocampal_R (40%) | 2.83 | 127 |
| | | Limbic | Amygdala_L (82%) | 2.56 | 51 |
| | | Limbic | Amygdala_R (77%) | 2.52 | 54 |
| | | Temporal | Temporal_Pole_Sup_L (38%) | 2.38 | 145 |
| | | Temporal | Temporal_Pole_Sup_R (31%) | 2.32 | 123 |
| | | Temporal | Temporal_Pole_Mid_L (44%) | 2.73 | 97 |
| | | Temporal | Temporal_Pole_Mid_R (39%) | 2.57 | 136 |
| | | Temporal | Temporal_Inf_L (50%) | 3.06 | 471 |
| | | Temporal | Temporal_Inf_R (40%) | 2.98 | 433 |
| | | Temporal | Temporal_Mid_L (20%) | 2.74 | 292 |
| | | Occipital | Fusiform_L (26%) | 3.11 | 181 |
| | | Occipital | Fusiform_R (45%) | 3.13 | 194 |
| **10** | Precuneus (85%) | Parietal | Parietal_Sup_L (50%) | 3.04 | 315 |
| | DMN (41%) | Parietal | Parietal_Sup_R (40%) | 3.05 | 257 |
| | Visuospatial (38%) | Parietal | Parietal_Inf_L (43%) | 2.62 | 299 |
| | | Parietal | Parietal_Inf_R (57%) | 2.92 | 240 |
| | | Parietal | Angular_L (31%) | 2.35 | 105 |
| | | Parietal | Angular_R (49%) | 2.91 | 241 |
| | | Parietal | Precuneus_L (65%) | 3.11 | 700 |
| | | Parietal | Precuneus_R (67%) | 3.22 | 629 |
| | | Parietal | Paracentral_Lobule_R (21%) | 2.28 | 47 |
| | | Occipital | Cuneus_L (46%) | 2.51 | 206 |
| | | Occipital | Cuneus_R (55%) | 2.70 | 239 |
| | | Occipital | Occipital_Sup_L (39%) | 2.74 | 156 |
| | | Occipital | Occipital_Sup_R (47%) | 3.14 | 203 |
| | | Occipital | Occipital_Mid_R (25%) | 2.76 | 149 |
| | | Limbic | Cingulum_Mid_L (25%) | 2.65 | 156 |
| | | Limbic | Cingulum_Mid_R (22%) | 2.61 | 135 |
| | | Limbic | Cingulum_Post_L (36%) | 2.38 | 49 |
| **11** | Primary visual (97%) | Occipital | Calcarine_L (65%) | 3.20 | 423 |
| | High visual (42%) | Occipital | Calcarine_R (82%) | 3.35 | 446 |
| | Precuneus (54%) | Occipital | Cuneus_L (85%) | 3.60 | 382 |
| | | Occipital | Cuneus_R (89%) | 3.71 | 387 |
| | | Occipital | Lingual_L (41%) | 2.56 | 270 |
| | | Occipital | Lingual_R (43%) | 2.50 | 294 |
| | | Occipital | Occipital_Sup_L (74%) | 3.29 | 292 |
| | | Occipital | Occipital_Sup_R (73%) | 3.29 | 313 |
| | | Occipital | Occipital_Mid_L (53%) | 2.74 | 511 |
| | | Occipital | Occipital_Mid_R (61%) | 2.97 | 361 |
| | | Parietal | Precuneus_R (23%) | 2.51 | 214 |
| **12** | Dorsal DMN (41%) | Frontal | Frontal_Sup_Orb_L (49%) | 2.77 | 143 |
| | | Frontal | Frontal_Sup_Orb_R (47%) | 2.72 | 146 |
| | | Frontal | Frontal_Mid_Orb_L (51%) | 2.92 | 139 |
| | | Frontal | Frontal_Mid_Orb_R (43%) | 2.90 | 127 |
| | | Frontal | Frontal_Inf_Tri_L (27%) | 2.54 | 193 |
| | | Frontal | Frontal_Inf_Tri_R (21%) | 2.47 | 133 |
| | | Frontal | Frontal_Inf_Orb_L (43%) | 2.66 | 215 |
| | | Frontal | Frontal_Inf_Orb_R (38%) | 2.75 | 193 |
| | | Frontal | Olfactory_L (32%) | 2.60 | 28 |
| | | Frontal | Olfactory_R (39%) | 2.45 | 32 |
| | | Frontal | Frontal_Sup_Medial_L (24%) | 3.03 | 200 |
| | | Frontal | Frontal_Sup_Medial_R (22%) | 3.09 | 144 |
| | | Frontal | Frontal_Sup_Orb_Medial_L (77%) | 3.55 | 174 |
| | | Frontal | Frontal_Sup_Orb_Medial_R (81%) | 2.64 | 212 |
| | | Frontal | Rectus_L (67%) | 2.64 | 174 |
| | | Frontal | Rectus_R (63%) | 2.63 | 128 |
| | | Limbic | Insula_L (21%) | 2.38 | 121 |

| | | | | | |
|---|---|---|---|---|---|
| | | Limbic | Insula_R (24%) | 2.37 | 131 |
| | | Limbic | Cingulum_Ant_L (64%) | 3.72 | 271 |
| | | Limbic | Cingulum_Ant_R (61%) | 3.26 | 241 |
| | | Subcortical | Caudate_L (48%) | 2.93 | 134 |
| | | Subcortical | Caudate_R (50%) | 2.88 | 144 |
| | | Subcortical | Putamen_L (26%) | 2.51 | 81 |
| | | Subcortical | Putamen_R (27%) | 2.55 | 87 |
| **13** | Cerebellum (51%) | Occipital | Fusiform_R (27%) | 2.36 | 204 |
| | | Cereb | Cerebellum_Crus1_L (52%) | 3.07 | 391 |
| | | Cereb | Cerebellum_Crus1_R (55%) | 3.23 | 434 |
| | | Cereb | Cerebellum_Crus2_L (43%) | 2.87 | 247 |
| | | Cereb | Cerebellum_Crus2_R (33%) | 3.07 | 202 |
| | | Cereb | Cerebellum_3_L (28%) | 2.09 | 11 |
| | | Cereb | Cerebellum_3_R (32%) | 2.43 | 20 |
| | | Cereb | Cerebellum_4_5_L (60%) | 2.71 | 216 |
| | | Cereb | Cerebellum_4_5_R (72%) | 2.83 | 175 |
| | | Cereb | Cerebellum_6_L (91%) | 3.22 | 470 |
| | | Cereb | Cerebellum_6_R (88%) | 3.53 | 473 |
| | | Cereb | Cerebellum_7b_L (36%) | 2.90 | 64 |
| | | Cereb | Cerebellum_7b_R (24%) | 2.70 | 39 |
| | | Cereb | Cerebellum_8_L (36%) | 2.83 | 201 |
| | | Cereb | Cerebellum_8_R (33%) | 3.01 | 223 |
| | | Cereb | Cerebellum_9_L (51%) | 2.77 | 129 |
| | | Cereb | Cerebellum_9_R (52%) | 2.85 | 127 |
| | | Vermis | Vermis_4_5 (58%) | 2.78 | 106 |
| | | Vermis | Vermis_6 (97%) | 3.52 | 102 |
| | | Vermis | Vermis_7 (100%) | 3.98 | 59 |
| | | Vermis | Vermis_8 (100%) | 3.74 | 67 |
| | | Vermis | Vermis_9 (98%) | 3.68 | 56 |
| | | Vermis | Vermis_10 (63%) | 2.66 | 19 |
| **14** | Anterior salience (52%) | Frontal | Frontal_Sup_L (44%) | 3.33 | 471 |
| | | Frontal | Frontal_Sup_R (45%) | 3.19 | 521 |
| | | Frontal | Frontal_Mid_L (31%) | 2.87 | 452 |
| | | Frontal | Frontal_Mid_R (33%) | 2.84 | 500 |
| | | Frontal | Supp_Motor_Area_L (71%) | 3.21 | 468 |
| | | Frontal | Supp_Motor_Area_R (59%) | 3.28 | 394 |
| | | Frontal | Frontal_Sup_Medial_L (39%) | 3.57 | 331 |
| | | Frontal | Frontal_Sup_Medial_R (40%) | 3.67 | 259 |
| | | Limbic | Cingulum_Mid_L (24%) | 2.58 | 150 |
| | | Limbic | Cingulum_Mid_R (33%) | 2.84 | 202 |
| **15** | Dorsal DMN (44%) | Frontal | Frontal_Sup_L (36%) | 3.43 | 389 |
| | Anterior salience (42%) | Frontal | Frontal_Sup_R (30%) | 3.33 | 343 |
| | | Frontal | Frontal_Mid_L (30%) | 3.08 | 436 |
| | | Frontal | Frontal_Mid_R (30%) | 2.80 | 460 |
| | | Frontal | Frontal_Inf_Tri_L (22%) | 2.33 | 163 |
| | | Frontal | Frontal_Sup_Medial_L (62%) | 3.70 | 527 |
| | | Frontal | Frontal_Sup_Medial_R (60%) | 3.40 | 383 |
| | | Limbic | Cingulum_Ant_L (79%) | 3.57 | 338 |
| | | Limbic | Cingulum_Ant_R (87%) | 3.76 | 346 |
| **16** | Cerebellum (44%) | Cereb | Cerebellum_Crus1_L (59%) | 2.47 | 445 |
| | ECN (27%) | Cereb | Cerebellum_Crus1_R (53%) | 2.59 | 425 |
| | | Cereb | Cerebellum_Crus2_L (78%) | 3.63 | 452 |
| | | Cereb | Cerebellum_Crus2_R (60%) | 3.63 | 372 |
| | | Cereb | Cerebellum_7b_L (71%) | 3.80 | 127 |
| | | Cereb | Cerebellum_7b_R (61%) | 3.87 | 98 |
| | | Cereb | Cerebellum_8_L (64%) | 3.23 | 353 |
| | | Cereb | Cerebellum_8_R (60%) | 3.33 | 407 |
| | | Cereb | Cerebellum_9_L (56%) | 2.83 | 142 |
| | | Cereb | Cerebellum_9_R (57%) | 2.87 | 138 |

| | | | | | |
|---|---|---|---|---|---|
| | | Vermis | Vermis_7 (71%) | 2.63 | 42 |
| | | Vermis | Vermis_8 (72%) | 3.60 | 48 |
| | | Vermis | Vermis_9 (65%) | 2.53 | 37 |
